# The Swiss national program for the surveillance of influenza A viruses in pigs and humans: genetic variability and zoonotic transmissions from 2010 – 2022

**DOI:** 10.1101/2025.01.28.24319114

**Authors:** J. Lechmann, A. Szelecsenyi, S. Bruhn, M. Harisberger, M. Wyler, C. Bachofen, D. Hadorn, K. Tobler, F. Krauer, C. Fraefel, A. R. Gonçalves Cabecinhas

## Abstract

Influenza A viruses (IAV) are likely candidates for pandemics. This report summarizes the results of the Swiss national program for surveillance of influenza viruses in pigs and transmissions to humans between 2010 and 2022. Challenges and optimization options in the program are discussed.

Nasal swabs or lung tissue samples from pigs with influenza-like signs (e.g. fever, cough) were screened by real-time RT-PCR for swine influenza virus (SIV) genomes, including that of the 2009 pandemic strain A(H1N1)pdm09; positive samples were subtyped for H1, N1, H3 and N2 by RT-PCR and Sanger sequencing. In parallel, humans with influenza-like symptoms and recent contact with diseased pigs were asked to self-sample themselves with a nasal swab. Human swabs were tested for IAV, and positive swabs further subtyped to identify potential cross-species transmission between swine and humans.

In the pigs, SIV was detected in 375 of 674 farm visits. H1N1 is the only subtype detected in Swiss pigs so far. The (H1N1)pdm09 strain (HA clade 1A) was only detected in seven out of 375 SIV positive farm visits. Phylogenetic analyses from partial hemagglutinin (HA) and neuraminidase (NA) genome sequences indicate that the remaining pigs were infected with the Eurasian avian lineage (HA clade 1C), which is predominant in swine in Europe. The Swiss H1N1 strains form distinct clusters within HA clades 1C.2.1 and 1C.2.2 and seem to evolve comparably slowly. Infection of humans with SIV was identified in five cases. Sequence analysis assigned the five viruses to the Eurasian avian lineage (C), clades 1C.2.1 and 1C.2.2. There was no evidence for sustained human-to-human transmission.

Although no critical IAV variants seem to have emerged so far in Switzerland, further surveillance of influenza viruses at the swine-human interface is of major importance.

## Introduction

Influenza A viruses (IAV, genus Alphainfluenzavirus) belong to the family of *Orthomyxoviridae*, a highly heterogenic group of viruses currently comprising nine genera (https://ictv.global/taxonomy, accessed August 6, 2024). They are pleomorphic enveloped viruses of 80 – 120nm in diameter with a segmented single-stranded RNA genome of negative polarity of approximately 10 – 13.5kb in size (Fig. 1).

**Figure 1:**
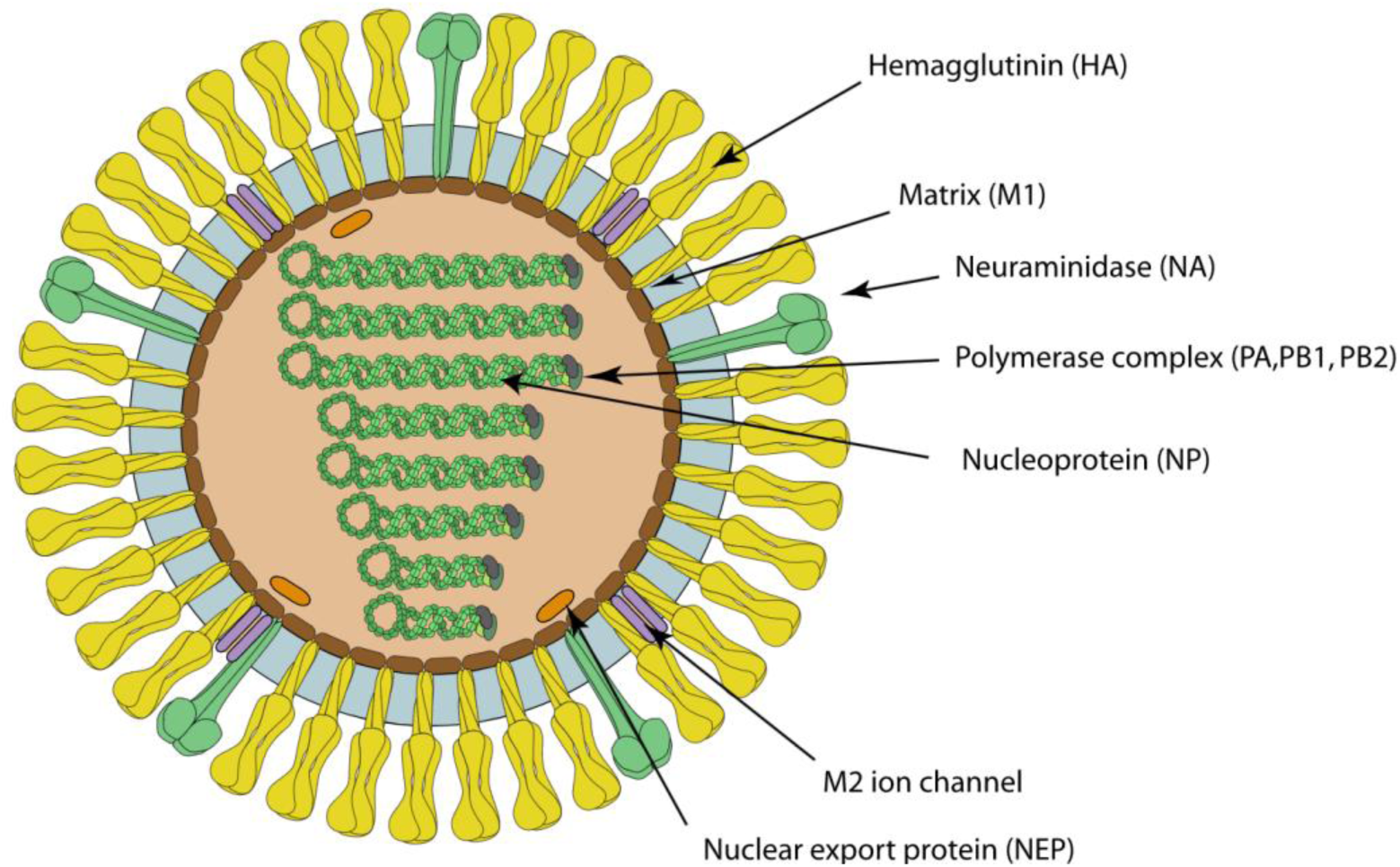
Influenza A virion structure (reprinted from ViralZone, SIB Swiss Institute of Bioinformatics, accessed July 28, 2023, https://viralzone.expasy.org).

IAVs are highly diverse and classified into subtypes, whose nomenclature is based on the genetic and antigenic properties of the two major surface glycoproteins hemagglutinin (HA) and neuraminidase (NA) (WHO, 1980). Three characteristics contribute to the rapid evolution and high diversity of IAV subtypes (Abdelwhab and Mettenleiter, 2023; Kessler et al., 2021; Krammer et al., 2018; Kuntz-Simon and Madec, 2009):

- The structural proteins HA and NA, which are responsible for attachment to the cell via the cellular receptor sialic acid (HA) and for mucus degradation in the upper respiratory tract as well as release of virus progeny from the cell after replication (NA), face strong evolutionary pressure.
- The RNA polymerase of influenza viruses lacks proofreading ability, conferring a more rapid accumulation of point mutations in the genome. In HA and NA, this process is referred to as *antigenic drift*.
- The segmented genome of IAV allows for reassortment, i.e. the reorganization of genome segments in cells co-infected with different IAVs, which may give rise to progeny viruses exhibiting markedly altered features. In HA and NA, this phenomenon is known as *antigenic shift*.

To date, 19 HA and 11 NA subtypes have been described (Karakus et al., 2024). With the exception of H17, H18, N10 and N11, which were recently detected in bats (Tong et al., 2012; Tong et al., 2013), all 17 HA and 9 NA subtypes have been detected in wild waterfowl (e.g. ducks, geese, gulls). In theory this translates into 144 different H/N combinations, of which at least 110 have been isolated to date (Pippig et al., 2016). Wild aquatic birds are therefore considered the natural reservoir of IAV (Bourret, 2018; Kessler et al., 2021). In the reservoir hosts, the virus establishes an enteric, usually asymptomatic infection and is shed via feces (Abdelwhab and Mettenleiter, 2023). Although the majority of these avian strains are host-specific, from wild waterfowl the virus can be transmitted to a wide range of other species including humans, horses, pigs, poultry, dogs, cats, ferrets, seals, mink via direct or indirect contact or aerosols (Fig 2). In most of these cross-species transmissions, viral fitness is insufficient in the new host, such that these variants undergo extinction after a few replication cycles. However, in rare cases the virus gradually adapts to the new host by accumulating mutations, allowing it to establish sustained transmission cycles within the species, species-specific lineages, and pathogenicity (Abdelwhab and Mettenleiter, 2023; Long et al., 2019).

**Figure 2:**
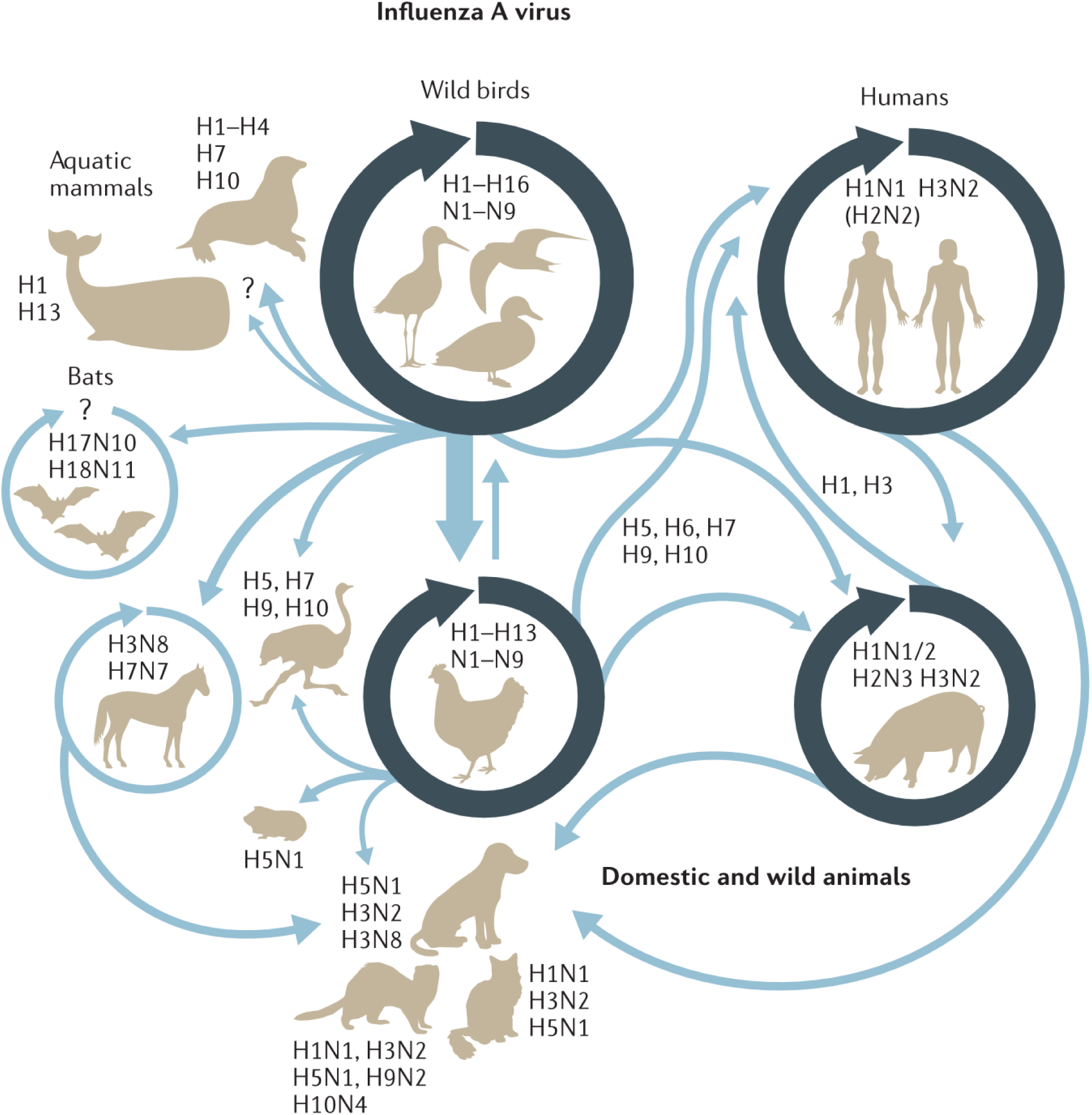
Interspecies transmission routes of IAV subtypes between different hosts. Subtypes from the wild bird reservoir can cross into many different species, sometimes via intermediate hosts and sometimes requiring adaptive mutations (light blue arrows). Specific subtypes predominate in certain species (dark blue circles) (adapted from Long et al., 2019). Having only recently been detected for the first time H5N1 infections in cows are missing in the figure (Eisfeld et al., 2024).

IAVs are circulating worldwide and have a high economic and health-related impact on humans and animals. In humans, pigs and horses, IAVs establish a respiratory infection with fever, headache, myalgia, lethargy, dry cough, sore throat, sneezing and nasal discharge (Kuntz-Simon and Madec, 2009; Vincent et al., 2020). Asymptomatic infection may also occur. The incubation period is between 1 and 3 days. Infection is usually self-limiting with recovery beginning 4 – 7 days after onset of clinical symptoms (Vincent et al., 2020). However, especially in young and immunocompromised humans and animals severe illness with secondary bacterial infections and pneumonia can develop and eventually lead to death (Abdelwhab and Mettenleiter, 2023; Krammer et al., 2018). The viruses are shed in nasal secretions and transmitted via direct contact between infected animals, indirect contact with virus-contaminated fomites, or through inhalation of aerosols (Nguyen et al., 2023). In swine, influenza is considered one of the most important pathogens of the respiratory disease complex, causing high economic losses due to poor performance and prolonged fattening periods (Ma, 2020).

In poultry and wild birds, a distinction is made between low (LPAI) and highly pathogenic avian influenza viruses (HPAI). While LPAI can cause mild disease, HPAI carry a specific mutation in the HA gene allowing it to spread systemically and cause a highly lethal disease not only affecting the respiratory, but also the gastrointestinal tract and the central nervous system of the birds (Taubenberger, 2019).

Continuous zoonotic and reverse zoonotic transmission of IAVs, especially between humans, swine and birds gives rise to endemic/enzootic strains but also to strains with pandemic potential (Abdelwhab and Mettenleiter, 2023; Kessler et al., 2021; Rajao et al., 2019). Endemic/enzootic IAV strains usually are the result of *antigenic drift* with subsequent minor changes in virus biology. These strains are constantly present in a confined region or population, at least partially recognized by the host’s immune system, and characterized by high morbidity (up to 100%) and low mortality (< 1%) (Vincent et al., 2020). The predominant endemic/enzootic IAVs currently circulating in pigs and humans are of the H1N1 and H3N2 subtypes (Fig. 2). However, porcine and human strains are antigenically and genetically distinct from each other (Krammer et al., 2018; Kuntz-Simon and Madec, 2009; Lewis et al., 2016). Pandemic strains, on the other hand, most often emerge upon *antigenic shift*, which is associated with abrupt changes in the antigenic properties of HA and NA. After infection with these strains, the host immune response is facing new epitopes, which are not or insufficiently covered by the immunological memory. Pandemic strains are characterized by rapid worldwide spread, high morbidity, more severe clinical signs, and potentially increased lethality (Krammer et al., 2018). Due to increasing immunity in the population and antigenic drift, pandemic strains may become endemic and replace the ones that previously circulated seasonally. The most recent example of strain replacement is the 2009 influenza H1N1 pandemic virus A(H1N1)pdm09 that replaced the former seasonal A(H1N1) in humans (Long et al., 2019). Since the beginning of the 20th century, humanity was struck by four IAV pandemics, three of which were of avian (H1N1 Spanish Flu 1918, H2N2 Asian Flu 1957 and H3N2 Hong Kong Flu 1968) and one of porcine origin (H1N1pdm09 2009) (Long et al., 2019, Saunders-Hastings and Krewski, 2016).

The cell receptor is a major determinant in host susceptibility to influenza viruses (Abdelwhab and Mettenleiter, 2023; Long et al., 2019). Owing to the location and density of cell receptors and the tropism of IAV, swine are considered a key species of concern in the emergence of pandemic strains (Bourret, 2018): human IAV strains have higher binding specificity to host glycoproteins containing sialic acid moieties in α2,6-galactose-linkage, which are most abundantly expressed in epithelial cells of the human upper respiratory tract. In contrast, avian strains preferentially bind to α2,3-galactose-linked sialic acid, which in avian species is present in the upper and the lower respiratory tract as well as the intestines, in humans, however, in the less accessible lower respiratory tract. Given the presence of both cell receptor types in pigs, it has long been believed that swine take on the role of a mixing vessel for viruses of human and avian origin, giving rise to novel reassorted variants to which humans are immunologically naive and highly susceptible. However, considering recent studies on the types and distribution of cell receptors in other species, the assumption that pigs are unique in expressing both receptor types and thus in being susceptible to infection by both avian and mammalian-adapted influenza strains no longer seems to be valid (Abdelwhab and Mettenleiter, 2023; de Graaf and Fouchier, 2014; Kuchipudi et al., 2021; Liu et al., 2023; Long et al., 2019). Instead, it is now hypothesized that additional factors determine receptor-specificity and pathogenesis, e.g. viral proteins NS1, polymerases, NA, and the host immune response (de Graaf and Fouchier, 2014) and that pigs serve as intermediate hosts in which pre-adaptation of avian subtypes to humans occurs (i.e., a gradual shift in IAV binding specificity for α2,3-gal to α2,6-gal sialic acid) thereby favoring transmission to humans (Bourret, 2018; Kuntz-Simon and Madec, 2009; Liu et al., 2023; Su et al., 2021). Thus, the pigs’ key role in IAV evolution is thought to be less related to physiological conditions than to husbandry practices where pigs and wild birds or poultry are kept in close proximity to each other, which increases the risk of interspecies transmissions (Long et al., 2019).

The risk of the emergence of novel influenza virus strains and global spread has risen significantly in recent decades due to the intensified livestock farming, invasion of wildlife habitats for agricultural use, and increasing transboundary movement of animals, humans, and goods (Kessler et al., 2021; Saunders-Hastings and Krewski, 2016). As a matter of fact, the first infections of HPAI H5N1 in dairy cattle have only recently been reported from the U.S.A. Interestingly, the virus replicates mainly in the mammary gland. Transmission between cattle is therefore thought to occur mainly through the milking process. From the cows, the infection has spilled over to humans, cats, rodents and poultry, highlighting the potential public health risk of this current outbreak (Eisfeld et al., 2024).

Given its potential role as a breeding ground and transmitter of new, potentially pandemic strains, surveillance of influenza viruses in pigs is of high importance. In the context of the 2009 pandemic caused by the originally porcine influenza virus A(H1N1)pdm09, the influenza virus surveillance in humans and pigs launched in 2001 in Switzerland was intensified. In collaboration with the Federal Food Safety and Veterinary Office (FSVO), the Federal Office of Public Health (FOPH), the Institute of Virology at the Vetsuisse Faculty of the University of Zurich, the National Reference Centre of Influenza (NRCI) at the University Hospital in Geneva and the Pig Health Service of SUISAG, a national program for surveillance of influenza viruses in pigs and in-contact humans was initiated, the aims of which were the following:

− Detection and molecular characterization of IAVs circulating in Swiss pigs and in-contact persons showing influenza-like symptoms, with a focus on HA and NA genes
− Where available, comparison of the genome sequences obtained from pigs and in-contact humans to identify potential cross-species transmission
− Early identification of the emergence of new IAV subtypes and changes in the epidemiologic situation in pigs

This study aims to present the results of the Swiss national program for the surveillance of influenza viruses in pigs and humans from 2010 – 2022, to discuss challenges in the success of national influenza virus surveillance and opportunities for optimization. The study does not claim to assess the prevalence of IAV in pigs.

## Material and methods

### Ethics statement

Porcine samples were taken by veterinarians (Pig Health Service, herd or private veterinarians) from clinically ill pigs and possibly contact animals with suspect SIV infection. The farms were located all over Switzerland.

The human samples from the target population (persons with influenza-like symptoms and recent contact with diseased pigs) were obtained by self-sampling. Based on an official letter from the Federal Office of Public Health (FOPH) stating that human data and samples were collected in accordance with the Swiss Epidemics Act (EpG SR 818.101), the cantonal ethics committee of Zurich waived ethical approval for this work (BASEC Req-2024-01630).

### Sampling of pigs and humans

Within the scope of the monitoring program, pig farmers were asked to report pigs with influenza-like signs such as cough and fever to the Pig Health Service. Nasal swabs (UTM Universal transport medium; Copan, Brescia, Italy) were then collected from acutely diseased pigs and contact animals by a veterinarian of the Pig Health Service, a herd veterinarian, or a private veterinarian. Persons with influenza-like symptoms and recent contact to these pigs were asked to swab themselves. Consenting individuals were provided with flexible minitip FLOQSwabs (Copan) and 3ml UTM tubes for self-sampling. In absence of recent influenza-like symptoms in humans, three (2010 – 2014) or two pigs (2015 – 2022) per farm were sampled.

Three pigs were sampled if in-contact humans were symptomatic. In addition to the specimens collected on-site, nasal swabs (UTM, Copan), lung tissue or dry lung swabs from pigs with suspected influenza infection were collected by institutes of veterinary pathology. Porcine samples were analyzed at the Institute of Virology, University of Zurich. Human samples were analyzed at the National Reference Centre of Influenza (NRCI) in Geneva. The samples were kept at 4°C (short term) or -20°C (long term) (Fig. 3).

**Figure 3:**
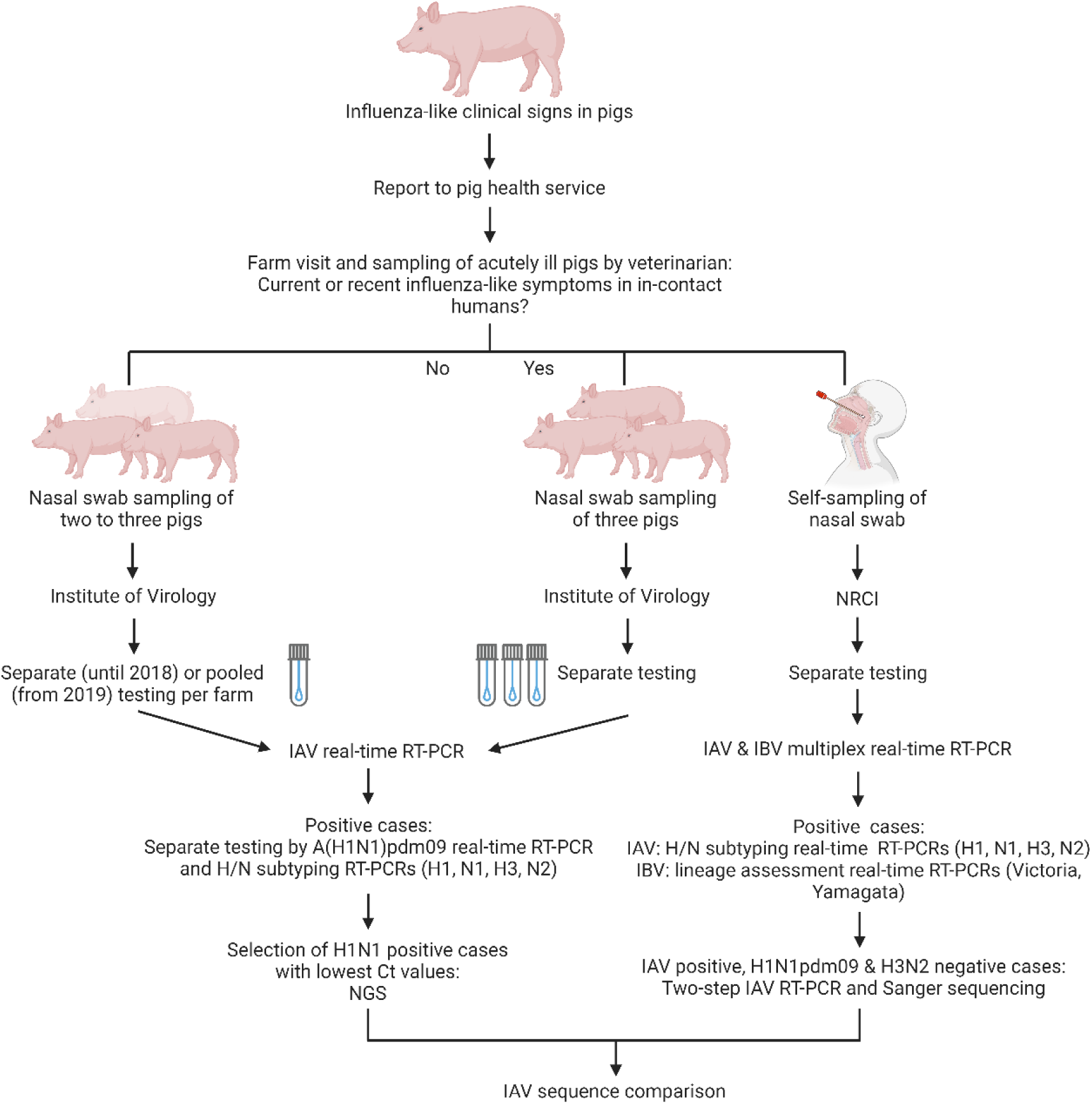
Workflow of sampling and testing of porcine and human nasal swab samples for IAV in the context of the Swiss national SIV monitoring program. A part of the porcine samples was subjected to Illumina whole-genome sequencing, the results of which will be published elsewhere. Human samples were additionally tested for Influenza B virus (IBV). (Figure created with BioRender.com)

### Testing of porcine samples

Until 2018, all porcine swabs were examined separately. From 2019, in case of absence of recent influenza-like symptoms in humans the samples were pooled, yielding a single sample per farm. If human samples were collected due to suspect influenza cases, the porcine samples on the respective farm were always analyzed individually (Fig. 3). The nasal swabs were thoroughly vortexed and centrifuged at 2671 x g (Heraeus Multifuge 3S-R, Thermo Scientific) for 10 min at room temperature (RT). The supernatant of each swab was mixed with antibiotic and antimycotic solution (100X, Sigma-Aldrich, St. Louis, U.S.). For pooling, 80 – 100µl of each supernatant were added to 560µl AVL buffer containing 5.6µl carrier RNA (QIAamp Viral RNA Mini Kit, Qiagen). The mixture was thoroughly vortexed and incubated for at least 10 min at RT. Lung tissue or lung swabs from institutes of veterinary pathology were pooled per farm. The dry swabs from lung tissue were immersed in 140µl PBS, thoroughly vortexed and centrifuged for 5 min at 16’000 x g. Lung tissue was subjected to three freeze-thaw cycles, centrifuged and 140µl of supernatant were used for RNA extraction. The supernatants of lung tissue and lung swabs were mixed with 560µl AVL buffer containing 5.6µl carrier RNA. Extraction was carried out according to the manufacturer’s instructions (QIAamp Viral RNA Mini Kit, Qiagen). If not processed immediately, the RNA was stored at -20°C or -80°C.

### Swine Influenza A real-time RT-PCR

For primary SIV detection, the extracted RNA was subjected to real-time RT-PCR targeting the matrix protein gene of IAV as well as the reference host gene GAPDH (Adiavet SIV Real Time, Adiagène-BioX Diagnostics, Ploufragan, France). Samples were tested in duplicate, using 20µl master mix and 5µl RNA. Extracted RNA from SIV positive field samples served as positive control, nuclease-free water and an extraction control without sample RNA served as negative control. RT-PCR was performed on a 7900HT Fast (Life Technologies) or a QuantStudio 7 Flex Real-Time PCR System (Applied Biosystems) using the following cycle protocol: 10 min at 45°C, 10 min at 95°C, 40 cycles of 15 sec at 95°C and 1 min at 60°C. Samples were considered SIV positive, if they yielded a Ct value ≤ 40 and if an increase in fluorescence signal was observed in the amplification and multicomponent plot.

### Influenza A(H1N1)pdm09 real-time RT-PCR

All SIV positive samples were tested in duplicate for the pandemic H1N1 strain from 2009 by a specific real-time RT-PCR. The RT-PCR mix consisted of 12.5µl 2x QuantiTect Probe RT-PCR Master Mix (QuantiTect Probe RT-PCR Kit, Qiagen), 1µg bovine serum albumin (1mg/ml; Albumin Bovine Serum Protease Free, VWR, Leuven, Belgium), 400nM of each primer, 200nM probe (Schulze et al., 2010), 0.5µl QuantiTect RT Mix and a top-up of 3.5µl nuclease-free water (Supplementary Table 1). Twenty microliters of the PCR mix were distributed onto a 96-well plate and supplemented with 5µl RNA. Positive and negative controls were included, and RT-PCR was performed and interpreted as described above, using the following cycling protocol: 30 min at 50°C, 15 min at 95°C, 40 cycles of 15 sec at 95°C and 30 sec at 60°C.

### H1N1 and H3N2 subtyping RT-PCRs and Sanger sequencing

For subtyping, SIV positive samples were subjected to conventional RT-PCR. First, specific primers for H1 and N1 were used (H1N1 is the only SIV subtype detected in Switzerland so far). All samples that tested negative against H1 or N1 were subsequently tested for H3 and/or N2. The 25µl RT-PCR mix consisted of 5µl 5x Qiagen OneStep RT-PCR buffer (Qiagen OneStep RT-PCR Kit, Qiagen), 400nM of each dNTP (dNTP Mix, 10mM each, Qiagen), 1µl Qiagen OneStep RT-PCR Enzyme Mix, 600nM of each primer (Chiapponi et al., 2003), 10µl nuclease-free water and 5µl RNA (Supplementary Table 1). All samples, including positive, negative and extraction controls were tested in duplicate on a FlexCycler2 (Analytik Jena, Jena, Germany).

The cycling protocol included an initial step of 30 min at 48°C, followed by 10 min at 95°C and 40 cycles of 1 min at 95°C, 1 min at 50°C and 1 min at 72°C, and a final elongation for 7 min at 72°C. PCR products were analyzed in a 1.5% or 2% agarose gel containing GelRed Nucleic Acid Stain (10’000X, Biotium, Hayward, CA, U.S.). Bands of the expected size were excised, and DNA extracted using the QIAquick Gel Extraction Kit (Qiagen). For Sanger sequencing (Microsynth, Balgach, Switzerland), the total DNA concentration was measured by spectrophotometry (Nanodrop, Thermo Fisher Scientific, Kloten, Switzerland), and approximately 45ng DNA were mixed with 20µM of the respective primer forward and reverse and topped-up with nuclease-free water to a final volume of 15µl. Sequences were analyzed with NCBI nucleotide BLAST (https://blast.ncbi.nlm.nih.gov; Altschul et al., 1990)

### Sequence analysis and generation of phylogenetic trees

Trimming of the ends resulted in HA and NA sequences of up to 392 and 457 nucleotides, respectively. Sanger sequencing traces were processed using *Tracy* (v0.7.6, Rausch et al., 2020) with default parameters. For samples with a single trace, the *basecall* module was used, whereas for samples with multiple traces and/or replicates, the *assemble* subcommand was applied. Subsequently, sequences for each segment were aligned using *MAFFT* (v7.310, Katoh et al., 2002) with the options *--reorder --adjustdirectionaccurately --op 5*. The resulting alignments were manually curated and trimmed. The phylogenetic relationships of the sequences were inferred using the maximum-likelihood nearest-neighbor interchange algorithm implemented in *FastTree2* (v2.1.10, Price et al., 2010). The trees were rerooted using the *root* command from the *ape* package (v5.8, Paradis et al., 2004) with AB628080/JX138509 and AB628082 used as roots for the HA and NA trees, respectively. Graphical representation of the trees was generated using ggtree (v3.12.0, Yu et al., 2017) and ggnewscale (v0.5.0 Campitelli, 2020).

Classification of HA sequences was performed using the online tool Subspecies Classification (with Swine Influenza H1 global classification, https://www.bv-brc.org/app/SubspeciesClassification, accessed August 4, 2024). To address limitations in NA segment classification, an in-house system was developed. Briefly, the aforementioned trimmed alignments were used to calculate a phylogenetic distance matrix using FAMSA (v2.2.2, Deorowicz et al., 2016) with the settings -square_matrix -dist_export. The dimensionality of the resulting distance matrix was reduced using the classical multidimensional scaling (MDS) method, implemented via the cmdscale function in R (v4.4.0, R Core Team, 2013). The visible segregation of sequences was ultimately used as an approximation for NA segment classification. All utilized scripts are available at https://github.com/mwylerCH/SwissSIV.

All assembled Sanger sequences were deposited on GenBank under the accession numbers PQ663270 – PQ663585 (Supplementary Table 2).

### Testing of human samples

Prior to nucleic acid extraction, 200 – 400µl of nasopharyngeal swab supernatant were spiked with 20µl of cell culture supernatant of canine distemper virus (Onderstepoort vaccine strain), which served as extraction control and to assess the presence of PCR inhibitors in the samples. Viral RNA was then extracted using Nuclisens easyMAG (2010 – 2017) and EMAG (since 2018) magnetic bead systems (BioMérieux, Italy) according to the manufacturer’s instructions. RNA was recovered in 25 – 50μl of NucliSens easyMAG Buffer 3. If not processed immediately, the RNA was stored at -80°C.

### Influenza A and B duplex real-time RT-PCR

For primary Influenza A (IAV) and B (IBV) detection, the extracted RNA was subjected to duplex real-time RT-PCR targeting the matrix protein of IAV and the non-structural protein of IBV viruses. Extracted RNA from IAV and IBV reference strains served as extraction and positive controls, and nuclease-free water (Promega, Wisconsin, U.S.) as negative control. The real-time RT-PCR mix consisted of 12.5μl of 2X Reaction Mix with ROX as passive reference (Invitrogen, California, U.S.), 0.5µl SuperScript III/Platinum Taq Mix (Invitrogen), 600nM of each primer, 200nM of each probe (adapted from CDC Realtime RTPCR (rRTPCR) Protocol for Detection and Characterization of Influenza (version 2007)), 4µl of nuclease-free water (Promega) and 5µl RNA (Supplementary Table 1). Real-time RT-PCR was performed on StepOnePlus and QuantStudio 5 Real-Time PCR Systems (Applied Biosystems) using the following cycling protocol: 30 min at 50°C, 2 min at 95°C, 45 cycles of 15 sec at 95°C, and 30 sec at 55°C. Samples were considered positive if they yielded a Ct value ≤ 40 and if in the amplification and multicomponent plots an increase in fluorescent signal was be observed.

### Influenza A subtyping and influenza B lineage assessment real-time RT-PCRs

Currently circulating human seasonal Influenza A subtypes are A(H1N1)pdm09 and A(H3N2). In parallel to IAV and IBV screening, all samples were subtyped using either H1pdm09 and H3 specific singleplex real-time RT-PCRs (2010 – 2017) or a quadruplex real-time RT-PCR targeting H1pdm09 and H3, as well as N1 and N2 genes (since 2018). Lineage (Yamagata and Victoria) was assessed for IBV positive samples (Fig. 3). Extracted RNA from Influenza A(H3N2), A(H1N1)pdm09, Influenza B Yamagata and Victoria reference strains served as positive controls, and nuclease-free water as negative control. The real-time RT-PCR mix consisted of 12.5μl of 2X Reaction Mix with ROX (Invitrogen), 0.5µl SuperScript III/Platinum Taq Mix (Invitrogen), 300 – 1200nM of target specific primers, 200 – 400nM of target specific probe (adapted from CDC Realtime RTPCR (rRTPCR) Protocol for Detection and Characterization of Influenza (Version 2007), Henritzi et al. (2016), Schweiger et al. (2000) and Watzinger et al. (2004)), 1µl of nuclease-free water, and 5µl RNA (Supplementary Table 1). Real-time RT-PCRs were performed as described above.

Suspected cases of SIV, i.e. IAV real-time RT-PCR positive or equivocal (after retesting), and H1N1 and H3N2 real-time RT-PCR negative samples were subjected to sequencing.

### RT-PCR and Sanger sequencing of samples suspected to be positive for zoonotic Influenza A

Zoonotic origin of the virus was considered probable when a positive IAV real-time RT-PCR (pan A) and negative H1N1pdm09 and H3N2 subtyping real-time RT-PCRs were observed. In these cases, a conventional RT-PCR using universal IAV primers was performed, and positive samples were subjected to Sanger sequencing. In case of low yield for the universal RT-PCR, a sequential PCR using segment-specific IAV primers was attempted. The porcine origin was confirmed if by Sanger sequencing a partial or full sequence of at least one viral segment of SIV was obtained.

In detail, the extracted RNA (12.5 µl) was reverse transcribed into cDNA. The reaction mix consisted of 10µl of 5X First-Strand Buffer (Invitrogen), 200U of SuperScript II Reverse Transcriptase (200U/µl; Invitrogen), 40U of Protector RNase inhibitor (40U/µl; Roche Diagnostics, Mannheim, Germany), 10mM dithiothreitol (0.1M; Invitrogen), 13.5µl nuclease-free water (Promega), 1.5mM dNTP (15mM; Amersham, Buckinghamshire, U.K.), and 1µg IAV universal primer (5’ – AGC AAA AGC AGG – 3’; 0.5µg/µl) (Hoffmann et al., 2001). Reverse transcription was performed at 42°C for 60 min, followed by 10 min at 95°C. The cDNA tubes were placed on ice and were either immediately processed for PCR or stored at -20°C.

The cDNA was used as template for amplification of the eight influenza virus genome segments using segment-specific primers (Supplementary Table 1) (Hoffmann et al., 2001). The PCR mix consisted of 5µl cDNA, 5µl 10X GeneAmp PCR buffer II (Applied Biosystems), 250nM of forward and reverse primers, 0.2mM dNTP (2.5mM; Amersham), 1% glycerol (Sigma-Aldrich, Steinheim, Germany), 1.5mM of GeneAmp MgCl_2_ (25mM; Applied Biosystems), 1.25µl AmpliTaq DNA Polymerase (5U/µl; Invitrogen), and 14.75µl nuclease-free water (Promega).

The first step of the amplification run consisted of a 4 min period at 94 °C and was followed by 35 cycles with the following conditions: 20 sec at 94°C, 30 sec at 58°C, and 7 min at 72°C. The program ended with one last cycle at 72 °C for 7 min.

For H1 and N1 genes, the resulting PCR products were sometimes subjected to additional segment-specific PCRs (i.e. nested PCR) to increase amplification yield (Supplementary Table 1). PCR mix conditions are comparable to those described above but in presence of 3mM of GeneAmp MgCl_2_ (25mM; Applied Biosystems). For H1, the amplification consisted of a 4 min period at 94 °C followed by 40 cycles with the following conditions: 15 sec at 94°C, 30 sec at 50°C, and 1 min at 72°C. The program ended with one last cycle at 72 °C for 5 min. For N1 gene, it consisted of one first cycle of 4 min at 94 °C, 30 sec at 50°C, and 2 min at 72°C, followed by 39 cycles of 30 sec at 94°C, 30 sec at 50°C, and 2 min at 72°C, followed by one cycle of 30 sec at 94°C, 30 sec at 50°C, and a final elongation for 10 min at 72°C.

Amplicons were purified using a MSB Spin PCRapace kit according to the manufacturer’s instructions (Invitek Molecular GmbH, Berlin, Germany) and sequenced using strain-specific primers. Sanger sequencing was performed on an ABI 3500xL Genetic Analyzer (Applied Biosystems, Singapore). Sequences were processed and stored in SmartGene’s Integrated Database Network System (IDNS) Influenza Module (https://apps.idns-smartgene.com/apps/Apps.po) and submitted to the global initiative on sharing all influenza data (GISAID) database (Shu and McCauley, 2017).

## Results

### Porcine samples

From 2010 to 2022, a total of 2303 cases of pigs with influenza-like signs were reported from approximately 1400 farms in Switzerland (Fig. 4). Over the years, the number of reports of farms with pigs showing influenza-like signs has continuously decreased from 320 in 2010 to 105 in 2022 (Fig. 5) (FSVO, 2023). The number of reports was evenly distributed over the entire area with pig husbandry (Fig. 4) and on average was highest in the last quarter of the year (Fig. 6).

**Figure 4:**
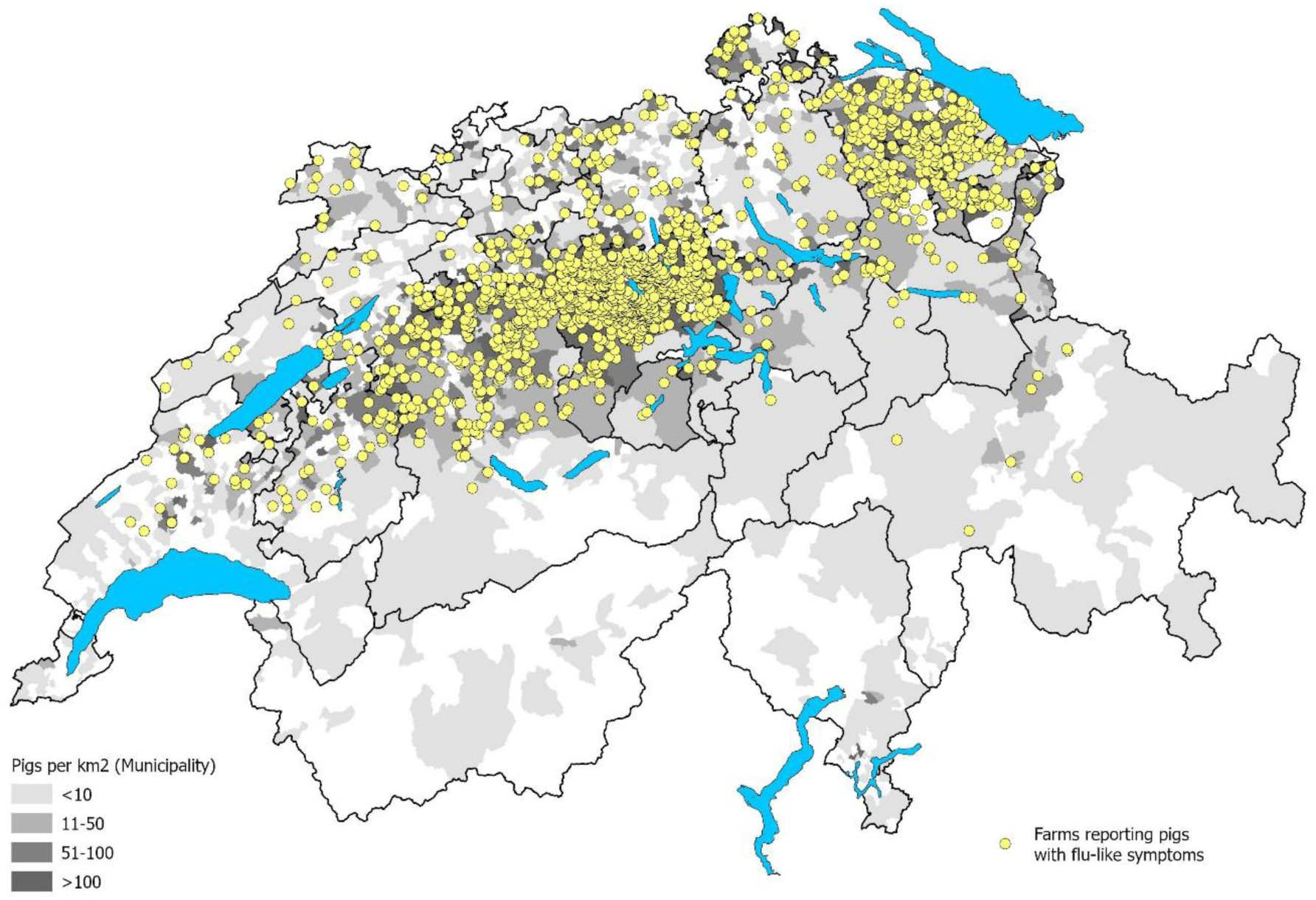
Regional distribution of farms reporting influenza-like signs in pigs from 2009 – 2022 (M. Binggeli, FSVO, 2023).

**Figure 5:**
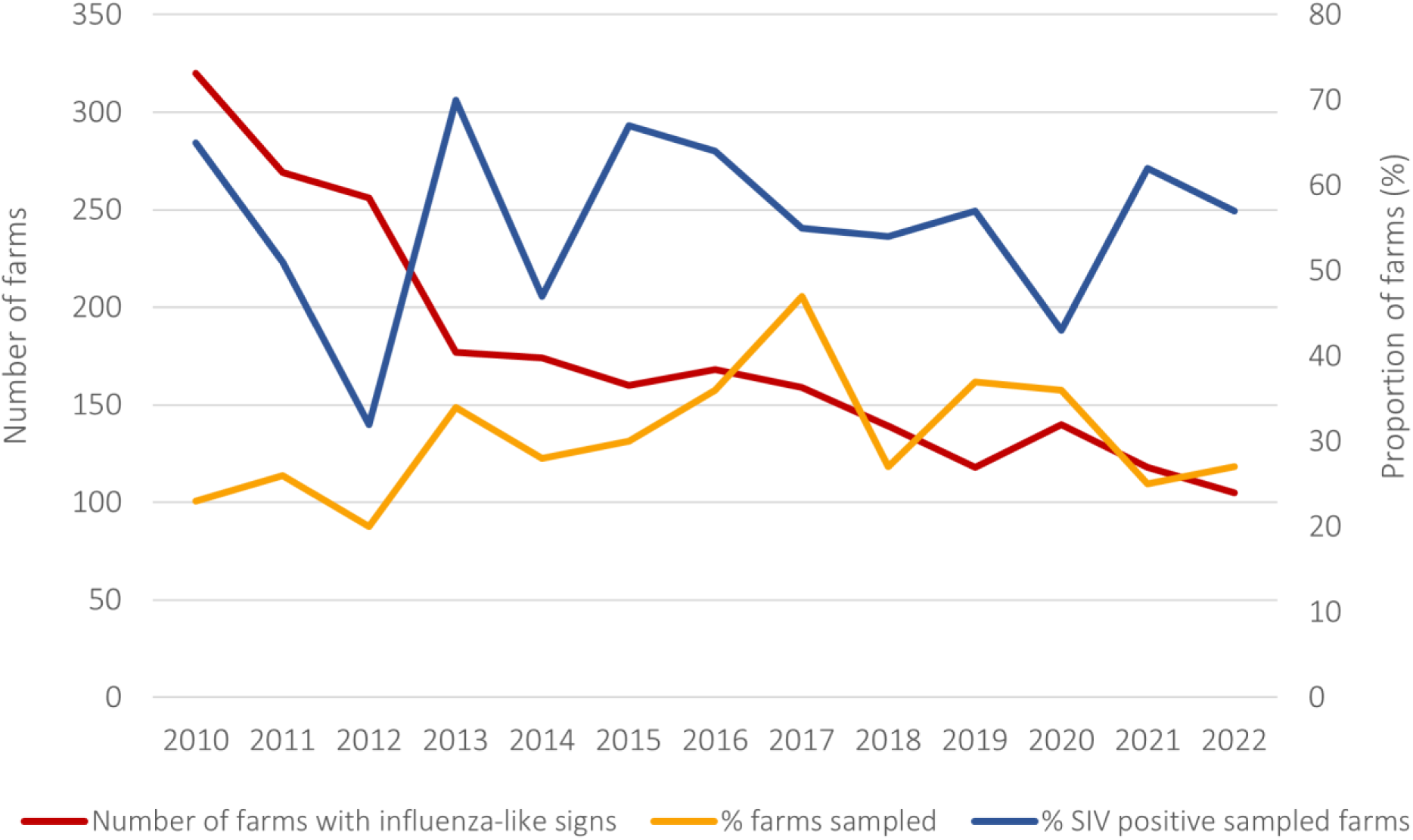
Number of farms reporting pigs with influenza-like signs (red line), proportion of farm visits with sampling (orange line) and proportion of sampled farms testing IAV positive (blue line) between 2010 and 2022.

**Figure 6:**
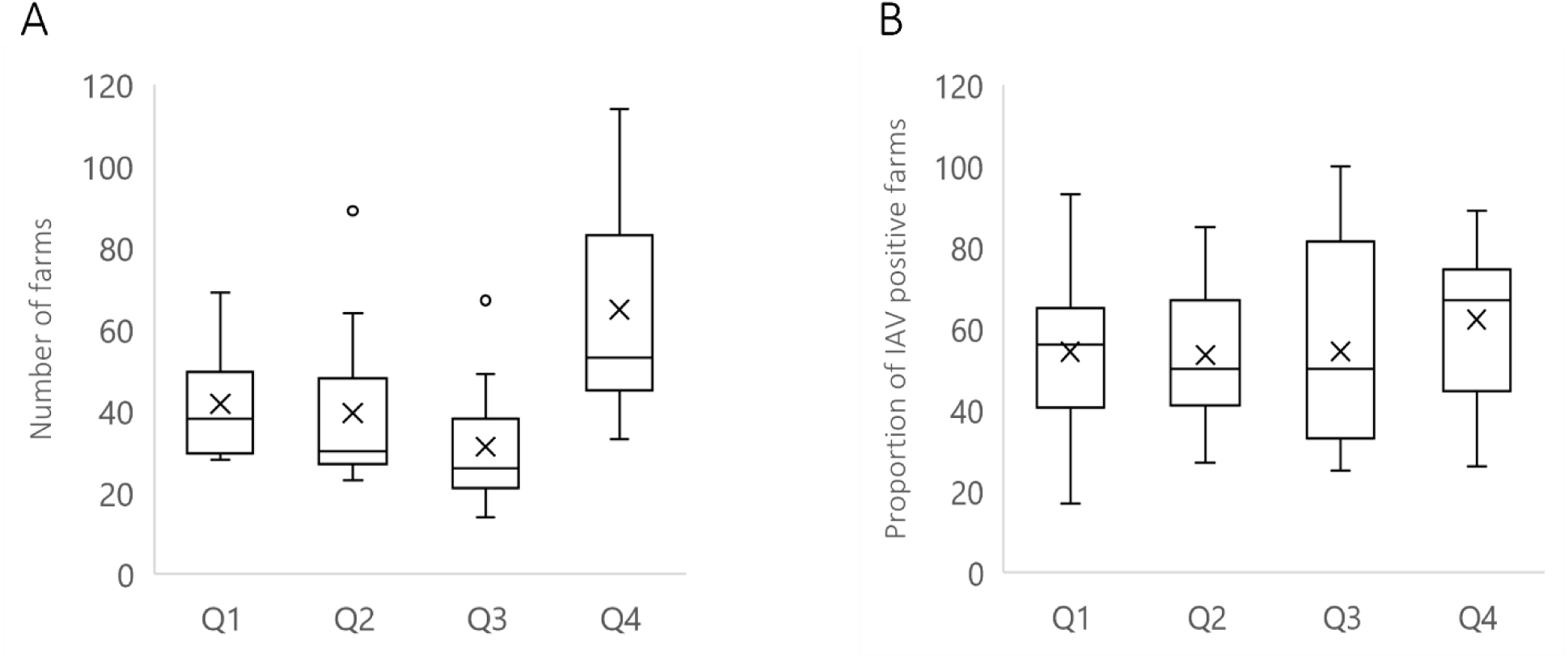
Number of farms reporting pigs with influenza-like signs (A), and percent of farm visits with IAV positive result by quarter (Q1 – Q4) (B) between 2010 and 2022. The X in each box corresponds to the mean value.

The proportion of farms sampled after reporting of influenza-like signs ranged from 20 – 47% (Fig. 5). On average 31% of farms were sampled per year. In total, 674 program-related farm visits took place between 2010 and 2022, in which 1660 porcine nasal swab samples were collected. In the laboratory, 145 pooled nasal swabs and 1515 unpooled nasal swabs were analyzed. In addition, in 107 cases lung material was analyzed. The percentage of farm visits (total n = 674) with positive IAV testing ranged between 32% and 70% (Fig. 5) and was highest in the last quarter of the year (Fig. 6). IAV was detected in a total of 375 (= 56%) farm visits.

The IAV detection rate was 44.94% (n = 746) in the nasal swab samples and 25.23% (n = 27) in the lung samples.

According to subtyping RT-PCR, none of the SIV positive samples could be assigned to H3N2 or H1N2 strains. Influenza A(H1N1)pdm09 was detected by specific real-time RT-PCR in four nasal swab samples in 2011 and three nasal swab samples in 2013. For one of these samples (SIV_75_13_H1) a Sanger sequence was obtained, confirming its type (Fig. 7). In the 375 IAV positive farm visits, the H1N1 subtype was detected on 196 occasions by subtyping RT-PCR and Sanger sequencing. In approximately 40 farm visits only H1, in approximately 80 farm visits only N1 and in approximately 60 farm visits no subtype was identified.

**Figure 7:**
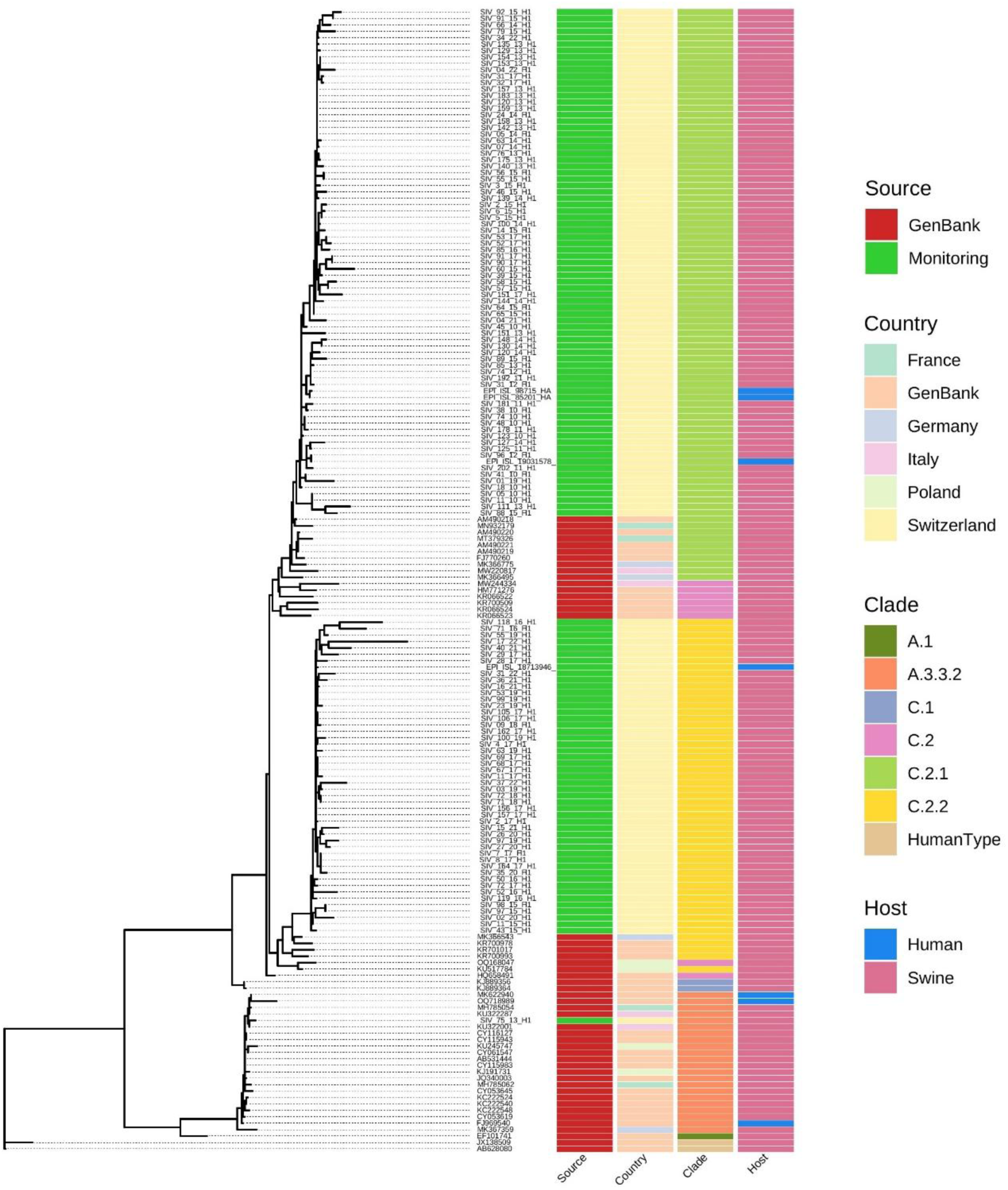
Maximum likelihood tree of the PCR-amplified region of the HA gene (approx. 450nt). The sequences obtained in the course of the Swiss national SIV monitoring program were labeled with ’SIV_number_year’. Sequences were characterized by country of origin, HA clade and host.

A total of 309 influenza sequences from PCR-amplified regions from pigs and humans were obtained after Sanger sequencing: 125 sequences from the HA segment and 184 from the NA segment. The HA sequences could be assigned to clades C.2.1 and C.2.2. The only exception is the one sample (SIV_75_13_H1) from 2013 mentioned above, which belongs to HA clade A.3.3.2 (Fig.7 and 9). Although clades C.2.1 and C.2.2 are also present in various European countries, the Swiss samples form distinct clusters that differ from strains in neighboring countries. This distinction is observed in both the HA segment (Fig. 7) and the NA segment (Fig. 8).

**Figure 8:**
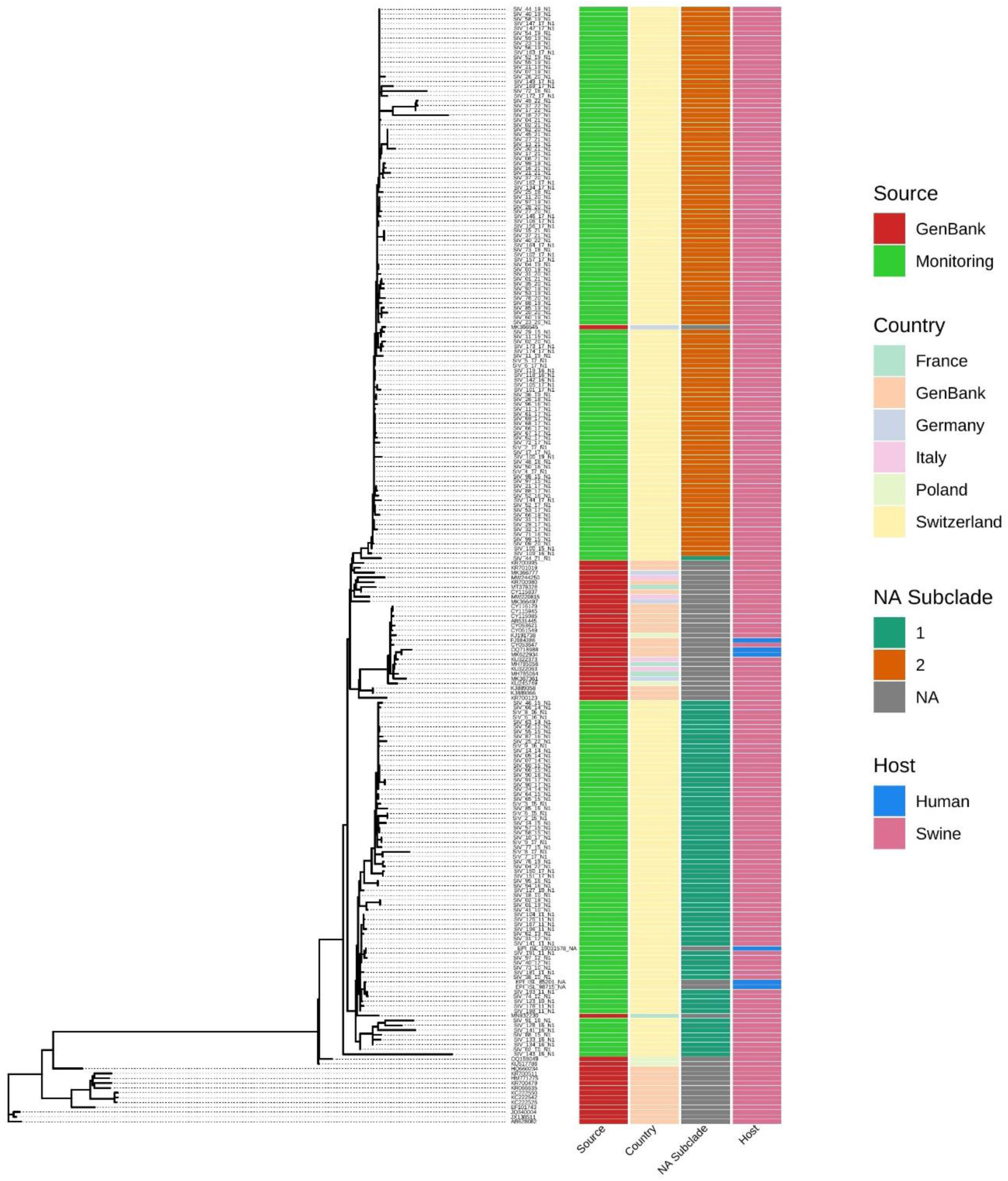
Maximum likelihood tree of the PCR-amplified region of the NA gene (approx. 450nt). The sequences obtained in the course of the Swiss national SIV monitoring program were labeled with ’SIV_number_year’. Sequences were characterized by country of origin, NA subclade and host.

Given the long-term nature of the national surveillance program, the temporal dynamics of the prevalence of SIV genotypes can be assessed (Fig. 9). The beginning of the program was characterized by the predominance of C.2.1 genotypes. Starting from 2015, the occurrence of this clade narrows, and the clade is superseded by clade C.2.2, however, without fully disappearing. By sequencing both HA and NA segments, potential reassortments could be investigated. Sequences for both segments were available for 78 samples (75 porcine samples, 3 human samples). Since canonical SIV subtyping into clades is typically performed on the HA gene sequence only, a custom method was developed to categorize the circulating NA genotypes in Switzerland. Similar to previous observations in the HA segment, the Swiss NA sequences also form two distinct clusters, here referred to as subclade 1 and subclade 2 (Fig. 8, Supplementary Fig. 1). Interestingly, NA subclade 1 tends to be combined with HA clade C.2.1, while subclade 2 is typically associated with C.2.2. However, six possible reassortments were identified (Fig. 9), in which a clade C.2.1 HA was combined with a subclade 2 NA and a clade C.2.2 HA with a subclade 1 NA.

**Figure 9:**
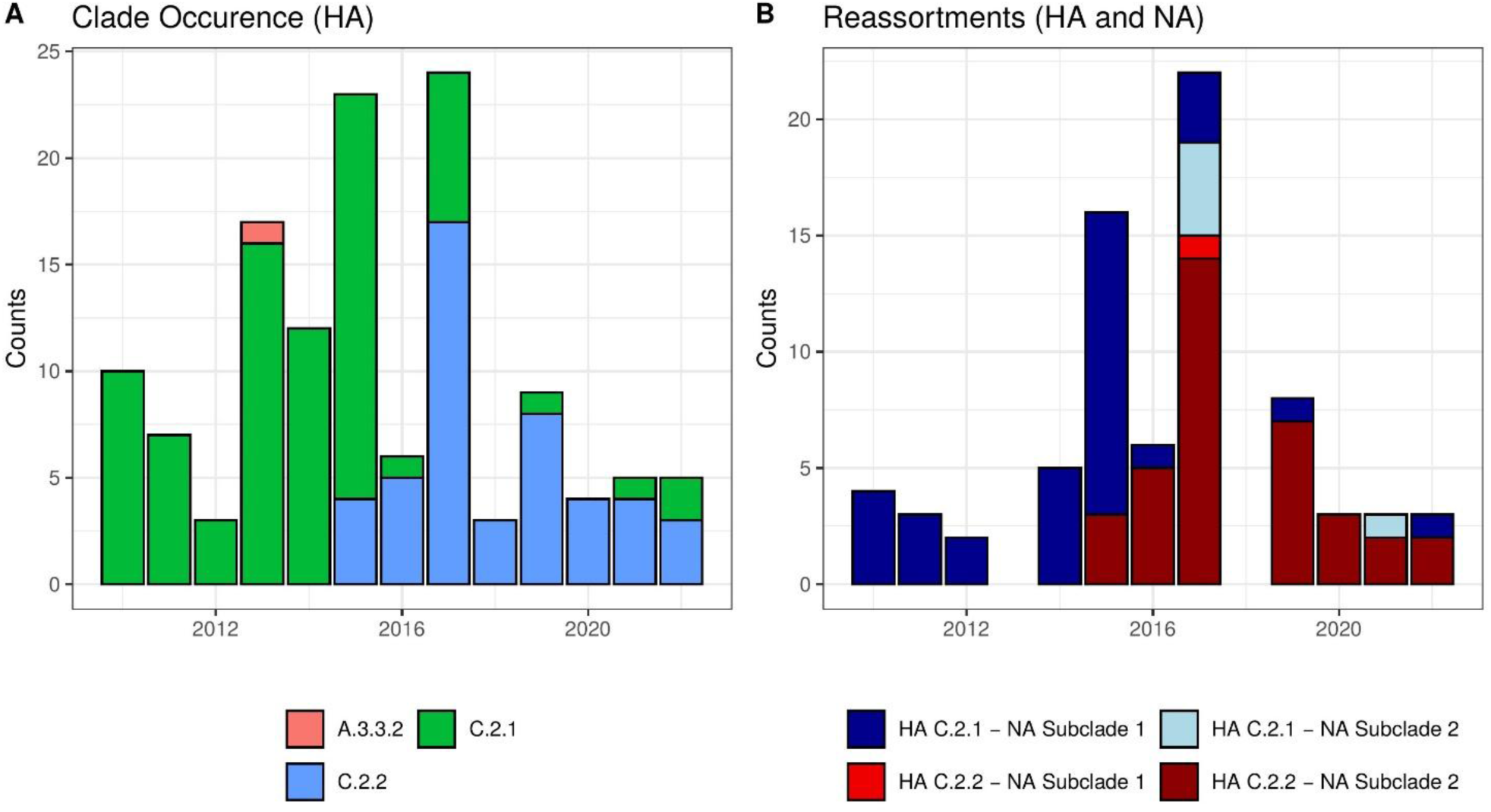
A) Temporal HA clade predominance of Swiss SIV strains from 2010 – 2022. B) Temporal overview of HA clade and NA subclade combinations with possible reassortments of Swiss SIV strains.

### Human samples

Between 2010 and 2022, in association with project-related farm visits (n = 674), a total of 150 humans on 118 farms reported influenza-like symptoms within ten days prior to the visit (Fig. 10) (FSVO, 2023). In 64 cases the onset of clinical symptoms was reported 1 – 3 days, in 86 cases 4 – 10 days before the farm visit. On average, 33% of humans with reported influenza-like symptoms were sampled per year. Over the years, the number of in-contact humans reporting symptoms has decreased from 22 in 2010 to 9 in 2022. In 455 project-related farm visits there were no reports of humans showing influenza-like symptoms in the 10 days prior to the farm visit, and in 103 visits, no information concerning illness in humans was available (FSVO, 2023).

**Figure 10:**
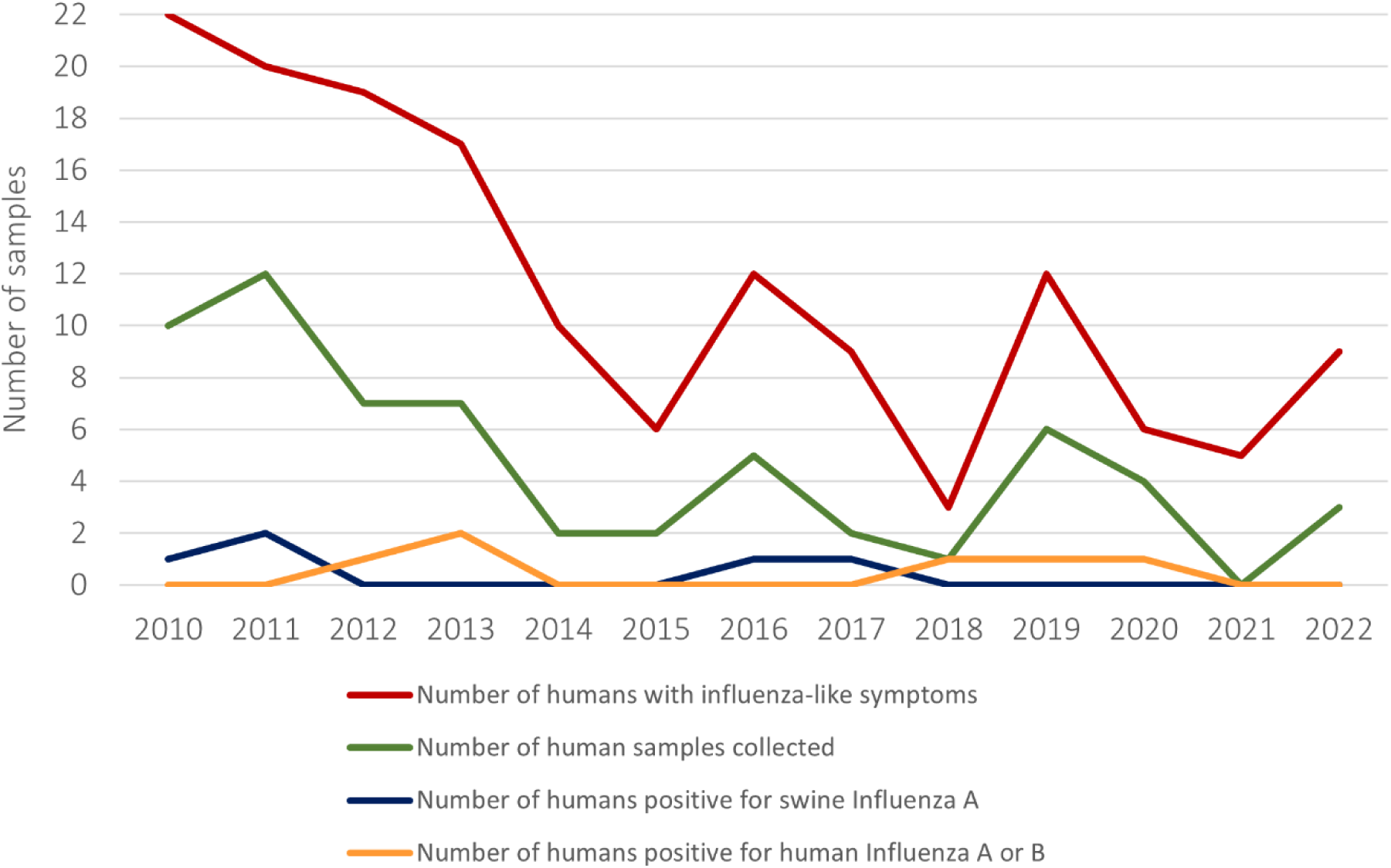
Number of humans reporting influenza-like symptoms in the context of the Swiss national SIV monitoring program (red line, n = 150), number of human samples collected (green line, n = 61), number of human samples positive for human Influenza A or B (orange line, n = 6) and number of human samples positive for SIV (blue line, n = 5) between 2010 and 2022.

Sixty-one individuals sent a self-collected nasal or nasopharyngeal swab to the NRCI (Fig. 10). Of these, 29 persons reported symptom onset 1 – 3 days, 10 persons 4 – 10 days, and six persons more than 10 days prior to the farm visit. In 16 cases no information on symptom onset was available. The most common symptoms reported were cough (60.6%), headache (36.1%), myalgia (27.9%), body temperature > 38°C, and rhinitis (18%).

Among the 61 sampled individuals, eleven tested positive for influenza virus. Three were positive for human influenza A (two A(H1N1)pdm09 and one A which could not be further characterized) and three for influenza B (two Yamagata and one Victoria) (Tab. 1). Five individuals were found positive for SIV (Tab. 2). Sequence analysis for HA and NA was successful in 4 and 3 cases, respectively, assigning the zoonotic viruses to the Eurasian avian lineage (clade HA-1C) (Fig. 7 and 8; Tab. 2).

**Table 1:** Human samples positive for human influenza A (IAV) and B virus (IBV).

| Date of collection | Symptom onset before sampling | Symptoms reported | Influenza Type |
| --- | --- | --- | --- |
| Dec 2012 | 1 – 3 days | Cough, headache, bronchitis | A(H1N1)pdm09 |
| Feb 2013 | 4 – 10 days | Headache, myalgia | A |
| Mar 2013 | 1 – 3 days | Fever, cough, headache, rhinitis | B Yamagata |
| Feb 2018 | 1 – 3 days | Fever, cough, headache, myalgia | B Yamagata |
| Apr 2019 | 4 – 10 days | Cough, headache, myalgia | A(H1N1)pdm09 |
| Jan 2020 | 4 – 10 days | Not reported | B Victoria |

**Table 2:** Human samples positive for SIV. GISAID = Global Initiative on Sharing All Influenza Data; HA = hemagglutinin; NA = neuraminidase; n.d. = not determinable.

| Sample collection | Symptoms | GISAID Accession | HA Clade | GenBank Accession HA | GenBank Accession NA |
| --- | --- | --- | --- | --- | --- |
| Oct 2010 | Fever, myalgia, rhinitis | EPI_ISL_85201 | C.2.1 | PQ663335 | PQ663568 |
| Apr 2011 | Cough, rhinitis | EPI_ISL_98715 | C.2.1 | PQ663336 | PQ663567 |
| Nov 2011 | Cough, rhinitis | EPI_ISL_19031578 | C.2.1 | PQ663346 | PQ663573 |
| Dec 2016 | Cough | EPI_ISL_242993 | n.d. |  |  |
| Dec 2017 | Cough, headache, myalgia, bronchitis | EPI_ISL_18713946 | C.2.2 | PQ663389 |  |

## Discussion

In the course of the Swiss national SIV monitoring program, reports of influenza-like signs in pigs and the detection of SIV in the respective farms occurred throughout the year, tending to be somewhat more frequent in the cold months. Nevertheless, as reported elsewhere, no seasonality of SIV can be inferred from these data (Fig. 6) (Anderson et al., 2021; Pippig et al., 2016; Vincent et al., 2020).

To date, three main subtypes (H1N1, H1N2, H3N2) including three major H1 lineages of SIV circulate in pigs in Europe which are distinct from their counterparts in North America and Asia (Anderson et al., 2016; Anderson et al., 2021; Bourret, 2018; Chiapponi et al., 2021; de Jong et al., 2007; Kessler et al., 2021; Kuntz-Simon and Madec, 2009; Lewis et al., 2016; Pippig et al., 2016; Vincent et al., 2020):

− The Eurasian avian lineage (H1avN1, clade HA-1C), which was transmitted from wild ducks to pigs and appeared in 1979
− The human-like H3N2 lineage, originating from the reassortment of human-like swine H3N2 and a H1avN1 in 1984
− The human seasonal lineage (H1huN2, clade HA-1B), which appeared in 1994 after reassortment of a human-like H3N2 and a seasonal human H1N1
− The classical swine lineage (H1N1pdm09, clade HA-1A), which circulates in the swine population since 2009, and is hypothesized to have arisen upon reassortment of H1avN1 and the North American triple reassortant H3N2

On farm level, H1avN1 is the dominant subtype in Europe, followed by other reassortant strains of H1N1 (including H1N1pdm09) and H1N2 subtypes, and H3N2 strains (Chiapponi et al., 2021).

In the present study, according to subtyping PCR none of the SIV positive samples could be assigned to H3N2 or H1N2 strains, making H1N1 the only subtype detected so far in the Swiss pig population (FSVO, 2023). In a small number of SIV positive samples, we found evidence for reverse zoonosis in the Swiss pig population: Influenza A(H1N1)pdm09 (HA-1A) was detected in a total of seven pig samples by specific real-time RT-PCR, i.e. four in 2011 and three in 2013. This virus emerged as a result of viral reassortment between two influenza lineages that had been circulating in pigs for years, i.e. a triple-reassortant circulating in North America and the Eurasian avian-like lineage (Smith et al., 2009). It was first detected in humans at the North American-Mexican border in March and April 2009 and, having acquired the ability for efficient human-to-human transmission, was declared a pandemic by the World Health Organization (WHO) by June 2009. By July the virus was reported from 122 countries (Kessler et al., 2021; Saunders-Hastings and Krewski, 2016) including Switzerland, where the first human case was detected in April 2009. It rapidly became the predominant IAV variant circulating in humans, with death rates fortunately remaining low (Thomas and Kaiser, 2010). From humans, the virus made its way back into pigs via reverse zoonosis (Rajao et al., 2019; Saunders-Hastings and Krewski, 2016), which is also the presumed route of transmission in the here described seven Swiss pig cases. Whereas formerly circulating human H1N1 strains were completely replaced by the pandemic variant, co-circulation of the pandemic strain with endemic viruses was observed in the pig population. However, at farm level H1avN1 (clade HA-1C) remained the predominant subtype in European pigs (Chiapponi et al., 2021; Pippig et al., 2016; Thomas and Kaiser, 2010; Zell et al., 2020).

With the exception of the seven A(H1N1)pdm09 (HA-1A) positive samples as per specific real-time RT-PCR, all HA and NA RNA segments sequenced in the present study could be assigned to the Eurasian avian lineage (HA-1C), which is widespread in pigs in Europe (Anderson et al., 2016). According to H1 and N1 sequence alignments, the Swiss H1N1 strains cluster into two HA clades, i.e. C.2.1 and C.2.2. Since 2015, the HA typing has shown a gradual shift from clade C.2.1 to C.2.2 (Fig. 9). However, C.2.1 sequences are still detected. Despite their evolutive capacities, phylogenetic analyses indicate that compared to other countries there is little genetic diversity and that the genome composition of influenza viruses in Swiss pigs remained fairly stable over the years (Fig. 7 – 9). Only six reassortments between H1 clades and N1 subclades were identified (Fig. 9). The absence of H1N2 and H3N2 subtypes, which are circulating in the rest of Europe, and the comparatively low diversity of H1N1 strains can be at least partly attributed to the Swiss pig production system, which is inherently isolated, influenced by low animal import levels and subject to strict biosecurity measures (Chiapponi et al., 2021; Lewis et al., 2016). Similar specific genetic clusters were observed for other viruses in Swiss pigs such as hepatitis E virus (Vonlanthen-Specker et al., 2021) and atypical porcine pestivirus (Kaufmann et al., 2019).

We also found evidence for spillover of influenza virus from pigs to humans: among the 11 influenza virus positive swabs of in-contact humans, six tested positive for human strains and five (= 45%) tested positive for SIV (Tab. 1 and 2). Sequence analyses assigned the SIVs to the Eurasian avian lineage of H1N1 and to the HA clade which at that time was most frequently detected in the pigs, i.e. C.2.1 and C.2.2 (Fig. 7 and 9). Given that the five individuals also had symptoms typical for influenza (Tab. 2), it is highly likely that pigs were the source of infection. Antibody testing of convalescent sera would have permitted more conclusive information but was not performed. Since the first well-documented reports in 1958 and 1974, human infections with SIVs (both H1N1 and H3N2) have continued to be reported sporadically worldwide.

Usually, these infections caused only mild disease resembling human seasonal influenza (Anderson et al., 2021; Freidl et al., 2014; Gregory et al., 2003; Kuntz-Simon and Madec, 2009; Rambo-Martin et al., 2020). Sustained human-to-human transmission rarely followed and has not been observed in the present study either. Compared to the number of people directly or indirectly involved in pig production, the number of SIV infections detected in humans is low. Yet, considering that in the frame of the Swiss national SIV program 45% of influenza positive human cases tested positive for SIV and that SIV infections are indistinguishable from seasonal human influenza, a high number of undetected cases must be expected. This hypothesis is supported by seroepidemiological studies in which higher seroprevalences of SIV were found in pig farmers and butchers than in people without occupational contacts to pigs (Borkenhagen et al., 2020; Krumbholz et al, 2014; Sun et al., 2020).

It is undisputed that national and international monitoring programs in animals and humans with internationally coordinated early warning systems constitute an important pillar in the surveillance of influenza viruses worldwide. However, after more than 15 years since its launch, the Swiss national monitoring program for SIV has brought to light various difficulties and hurdles that need to be addressed in the future: (1) The number of farms reporting pigs with influenza-like signs has continuously decreased from 320 in 2010 to 105 in 2022 (Fig. 5) FSVO, 2023). Likewise, the number of in-contact humans reporting influenza-like symptoms has decreased from 22 to 9 over the same period (Fig. 10). In 103 cases, no information concerning illness in humans was available. These reduced numbers most likely do not reflect a decrease in infections but rather a decline in disease awareness after the 2009 pandemic. In addition, due to the endemic circulation of SIVs and the resulting herd immunity, severe symptoms associated with a high viral load are probably rather rare and mainly seen in co-infections with other pathogens. For the future, all stakeholders including the farmers, veterinarians, meat producers and veterinary authorities should take measures to increase disease awareness. (2) After notification of influenza-like symptoms, the rate of farms and individuals sampled remained at a more or less constant low level of approximately 30%. Well-orchestrated reporting, communication and cooperation between stakeholders is required in order to consistently sample pigs on reported farms. (3) In the IAV-positive farm visits, the rate of samples in which both HA and NA were successfully subtyped was rather low (196/375 = 52%). In approximately 11% only H1, in 21% only N1 and in 16% of cases no subtype was identified. For a large proportion of IAV positive samples, including human swabs, the Ct value after IAV real-time RT-PCR was rather high, making successful subtyping and sequencing more difficult. High Ct values can result from various causes: low viral loads in the sample material, suboptimal sampling, poor sample quality due to contamination with dirt and bacteria, improper sample handling and/or storage, or low-performing assays. In many cases, the clinical signs may only be recognized late, or the nasal swabs may be taken late in the infection cycle, i.e. at a time when maximum virus excretion is already over and virus clearance almost complete. Efficient SIV surveillance programs depend on sensitive and specific diagnostic methods which allow for cost-effective large-scale analysis. To detect circulating strains and new variants, currently several PCR assays are needed, which make the process expensive and time-consuming. Novel methods for high-throughput whole genome sequencing such as Illumina or Nanopore technologies, which have been extensively developed in recent years and have become more sensitive and affordable, could improve the surveillance of SIV and reduce the cost of virus subtyping in the long term (Goecke et al., 2018; Rambo-Martin et al., 2020). From a laboratory perspective, a future main goal of the Swiss national surveillance program is the implementation of a SIV MinION Nanopore sequencing protocol (King et al., 2020), that in parallel to the established Illumina whole-genome sequencing protocol (Kubacki et al., 2021) allows for cost-efficient untargeted sequencing of all influenza virus genome segments present in a sample.

## Data Availability

All data produced in the present study are available upon reasonable request to the authors.

https://github.com/mwylerCH/SwissSIV

https://apps.idns-smartgene.com/apps/Apps.po

https://gisaid.org/

## Acknowledgements

We would like to express our sincere appreciation to the farmers and veterinarians participating in the national SIV surveillance program. They are the key players in the success of the program and their collaboration makes an important contribution to tracking influenza virus evolution in Switzerland.

Many thanks to M. Binggeli (FSVO) for providing the geographical distribution on farms reporting coughing pigs.

