## Supplementary figures and images for "The Swiss national program for the surveillance of influenza A viruses in pigs and humans: genetic variability and zoonotic transmissions from 2010 – 2022"

### Supplementary Fig. 1

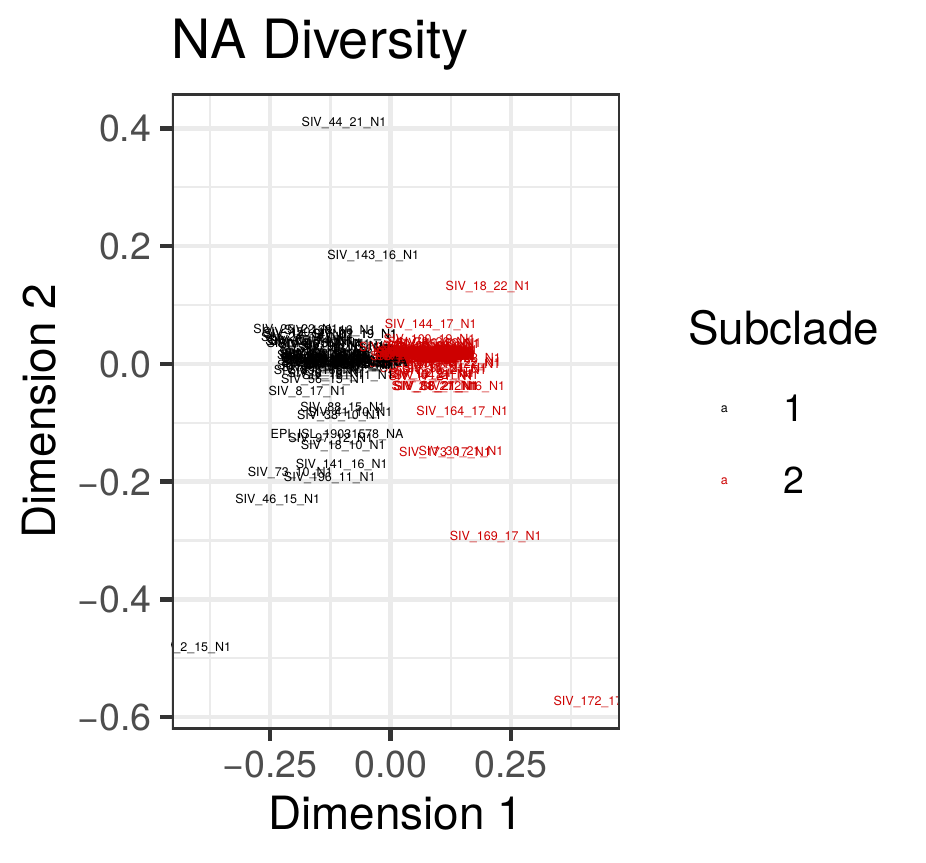
